# Intensive Versus Conservative Blood Pressure Lowering after Endovascular Therapy in Stroke: a meta-analysis of randomized controlled trials

**DOI:** 10.1101/2024.04.04.24305237

**Authors:** Ahmed Naji Mansoor, Vatsalya Choudhary, Zain Mohammad Nasser, Muskan Jain, Dhruvikumari Dayanand Sharma, Mateo Jaramillo Villegas, Sujaritha Janarthanam, Muhammad Ayyan, Simran Ravindra Nimal, Huzaifa Ahmad Cheema, Muhammad Ehsan, Muhammad Aemaz Ur Rehman, Sourbha S. Dani

**Affiliations:** Department of Medicine, Cardiff University School of Medicine, Cardiff, United Kingdom; Department of Medicine, Kasturba Medical College, Manipal, India; Royal Lancaster Infirmary, United Kingdom; Department of Medicine, Avalon University School of Medicine, Curaçao; Universidad CES, Medellin, Colombia; Department of Medicine, Sri Ramachandra University, India; Department of Medicine, King Edward Medical College, Pakistan; Department of Medicine, B.J. Medical College, Pune, India; Department of Neurology, University of Alabama; Department of Cardiology, Lahey Hospital and Medical Center, Burlington, MA

**Author notes:** **Corresponding Author:** Muhammad Ayyan, Address: Department of Medicine, King Edward Medical University, Nila Gumbad Chowk, Neela Gumbad Lahore, Punjab, Pakistan 54000, Twitter handle: @ayyan_77.

**Keywords:** meta-analysis, stroke, endovascular therapy

## Abstract

**Background:** The optimum systolic blood pressure after endovascular thrombectomy for acute ischemic stroke is uncertain. We aimed to perform an updated meta-analysis of randomized controlled trials to evaluate the safety and efficacy of more intensive blood pressure management as compared to less intensive blood pressure management.

**Methods:** We searched various electronic databases including Embase, MEDLINE (via PubMed), and CENTRAL to retrieve relevant randomized controlled trials (RCTs) on the clinical effects of more intensive blood pressure management after endovascular thrombectomy as compared to the less intensive management. We assessed the risk of bias using the revised Cochrane “Risk of bias” tool for randomized trials (RoB 2.0), calculated risk ratio (RR) with 95% confidence intervals (CI) for dichotomous outcomes, and mean difference (MD) with 95% CI for continuous outcomes.

**Results:** Our meta-analysis included 4 RCTs with a total of 1560 patients. According to our analysis, more intensive blood pressure management was associated with a statistically significant decrease in the number of patients showing functional independence (modified Rankin scale mRS score=0-2) at 90 days (RR 0.81; CI = 0.72-0.91; I^2^ = 12%). Regarding 90-day mortality, our pooled results from three RCTs showed no statistically significant difference between the more intensive blood pressure management group and the less intensive blood pressure management group (RR 1.17; CI = 0.90-1.52; I^2^ = 0%). There was no statistically significant difference between the two groups regarding the incidence of intracerebral hemorrhage (ICH) (RR 1.05; CI = 0.90-1.23; I^2^ = 0%) and the incidence of symptomatic intracerebral hemorrhage (sICH) (RR 1.10; CI = 0.76-1.60; I^2^ = 0%).

**Conclusion:** According to our meta-analysis, no benefit of intensive lowering of blood pressure was observed in terms of functional independence at 90 days, mortality rates, and incidence of intracerebral hemorrhage. Future large-scale trials should focus on other interventions to improve prognosis in these patients.

## Introduction

An Acute Ischemic Stroke (AIS) is an episode of sudden neurological dysfunction resulting from brain ischemia, which is associated with acute infarction on brain imaging ^1^. In the United States, AIS affects approximately 700,000 individuals annually and is responsible for over 150,000 deaths. AISs carry significant complications for patients, including depression, cognitive impairment, and disability, in addition to placing considerable financial burdens on healthcare systems ^2^.

Presently, endovascular thrombectomy (EVT) is a well-established and standard therapeutic approach for AIS resulting from a large vessel occlusion (LVO) ^3^. EVT is highly effective, with successful recanalization in 4 out of 5 procedures. In early intervention EVT, for every 2-3 patients treated, one extra patient attains a reduction in disability by at least one point on the modified Rankin Scale (mRS) ^4^. Favorable outcomes of EVT treatment are time-dependent: every hour delay from stroke onset to EVT initiation was linked to a 5% reduction in post-treatment functional independence. More recent trials suggest that carefully selected patients, based on initial infarct volume, have more favorable outcomes up to 24 hours post-symptom onset than standard medical therapy ^5,6^.

Recent advances in newer-generation thrombectomy devices, more efficient pretreatment admission processes, and more strict selection criteria for eligible patients have remarkably increased the efficacy and outcomes of EVT. In post-treatment care, observational studies have explored the effect of post-EVT blood pressure (BP) levels on AIS prognosis and treatment outcomes ^7–12^. Optimal blood pressure control is a challenging target, considering the adverse effects of both low and high blood pressure. On the one hand, higher blood pressure after EVT was associated with an increased risk of unfavorable safety outcomes in the form of sICH, mortality, and requiring hemicraniectomy. Blood pressure lower than 160/90 mm/Hg was associated with better 3-month functional independence rates than permissive BP management ^7,9^. On the other hand, very intensive BP lowering may compromise cerebral perfusion and increase the ischemic core ^13^.

Recent guidelines recommend a BP goal of <180/105 mmHg post successful reperfusion ^14^. Yet, a survey of American acute stroke centers revealed that there is no consensus on this between clinicians as most institutions do not have a standardized, post-treatment BP target. 36%, 28%, and 21% of institutions reported systolic blood pressure (SBP) targets of 120-139, 140-159, and ≤ 180 mmHg post-successful reperfusion, respectively ^15^._J Considering the absence of precise demonstrated guidelines and some evidence suggesting beneficial outcomes for intensive BP control, a previous meta-analysis was conducted on randomized clinical trials (RCTs) and observational studies that explored intensive BP monitoring for AIS patients post-EVT ^16^. However, it suggested insignificant benefits of intensive BP control. Two new RCTs have recently been published on this topic ^17,18^. Hence, this meta-analysis provides an updated review based on RCTs to compare and determine the efficacy of less and more intensive BP control post EVT._J

## Methods

This systematic review and meta-analysis, registered in Prospero with registration number CRD42023492018, was performed according to the guidelines of the Cochrane Handbook for Systematic Reviews of Interventions ^19^ and reported according to the Preferred Reporting Items for Systematic Reviews and Meta-Analysis (PRISMA) statement ^20^.

### Eligibility Criteria

#### Inclusion Criteria

Included in the analysis were studies meeting the following criteria: (1) Enrolment of individuals with acute ischemic stroke. (2) Individuals underwent endovascular thrombectomy with successful reperfusion, as defined by a modified Thrombolysis In Cerebral Ischemia (mTICI) score of ≥ 2b. (3) More Intensive blood pressure lowering as the intervention and conservative blood pressure lowering as the comparator. (4) Assessment of outcomes such as 90-day modified Rankin Scale (mRs) score, 90-day mortality, symptomatic intracranial hemorrhage (sICH), or mortality. (5) Employing a randomized controlled trial (RCT), study design.

#### Exclusion Criteria

All study types other than RCTs, and those that did not measure our outcomes were excluded. Information sources and search strategy

To comprehensively gather relevant studies, we searched critical electronic databases such as the Cochrane Central Register of Controlled Trials (CENTRAL), MEDLINE (via PubMed), Embase (via Ovid), and ClinicalTrials.gov. Grey literature sources like ProQuest and OpenGrey were also used to find relevant RCTs. Reference lists of included articles and relevant reviews were searched to ensure a comprehensive search. Different keywords like “ischemic stroke,” “endovascular thrombectomy,” and “blood pressure management” were used to search for relevant RCTs.

#### Outcome Measures

The primary outcome of interest is the incidence of 90-day modified Rankin Scale (mRS) scores ranging from 0 to 2. Additionally, secondary outcomes include: 1) incidence of 90-day mortality, 2) any intracerebral hemorrhage (ICH), and 3) symptomatic ICH (sICH) (intracerebral hemorrhage associated with ≥ 4 points increase from the baseline NIH Stroke Scale [NIHSS] score) ^21^.

#### Study Selection and data extraction

All the articles retrieved from the search were imported into Mendeley Desktop 1.19.8. After deduplication, two reviewers carried out the screening process. Screening was done, first according to the title and abstract of the studies and then by reading the full text of the articles. Any disagreements during full-text screening were resolved through discussion; in some cases, a third reviewer acted as an arbiter. The study selection process has been represented via a PRISMA flowchart (Figure 1).

**Figure 1.**
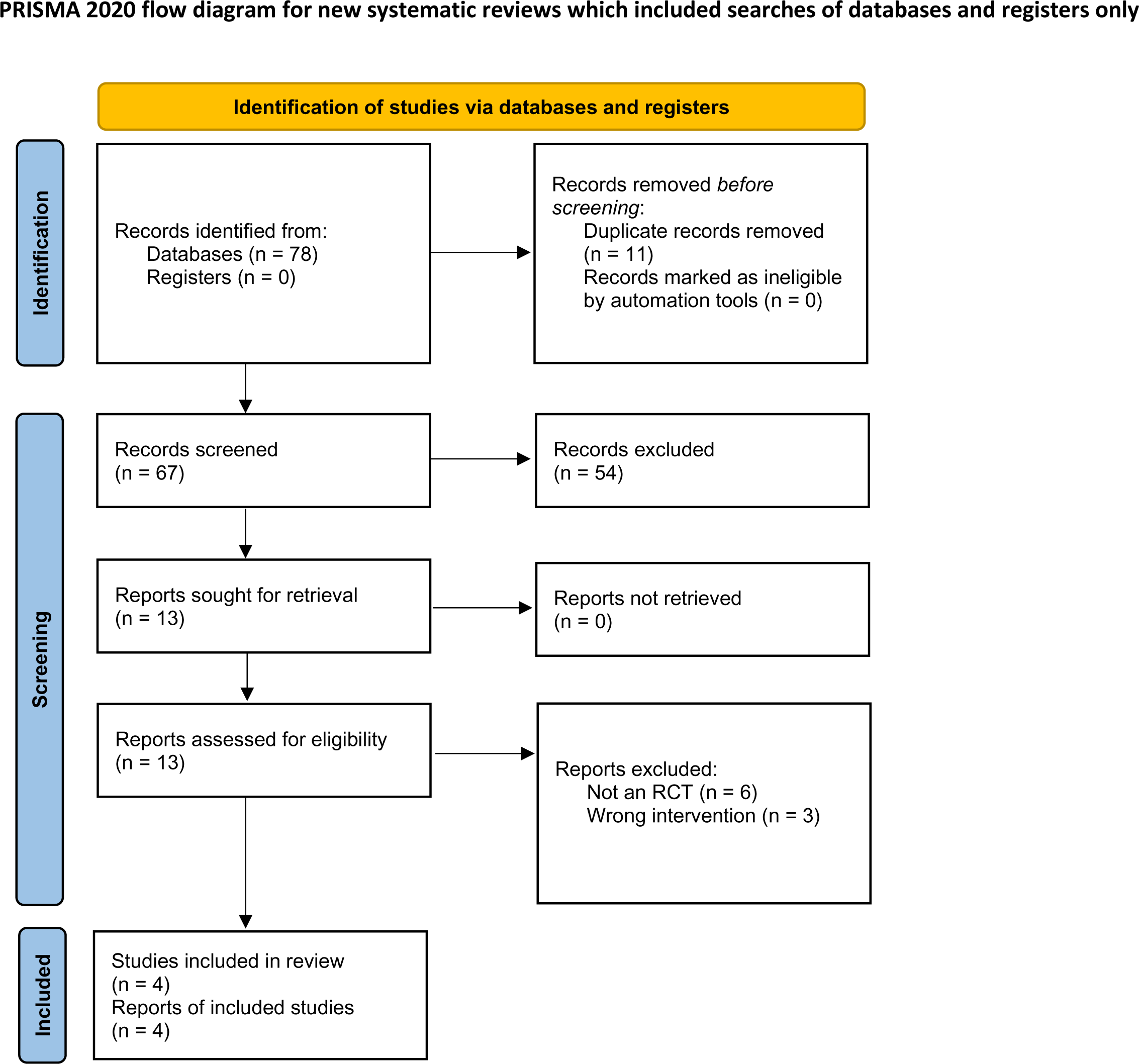
PRISMA 2020 flow chart. Flow chart of included and excluded trials. PRISMA, Preferred Reporting Items for Systematic Reviews and Meta-Analyses. *From:* Page MJ, McKenzie JE, Bossuyt PM, Boutron I, Hoffmann TC, Mulrow CD, et al. The PRISMA 2020 statement: an updated guideline for reporting systematic reviews. BMJ 2021;372:n71. doi: 10.1136/bmj.n71 For more information, visit: http://www.prisma-statement.org/

**Figure 2.**
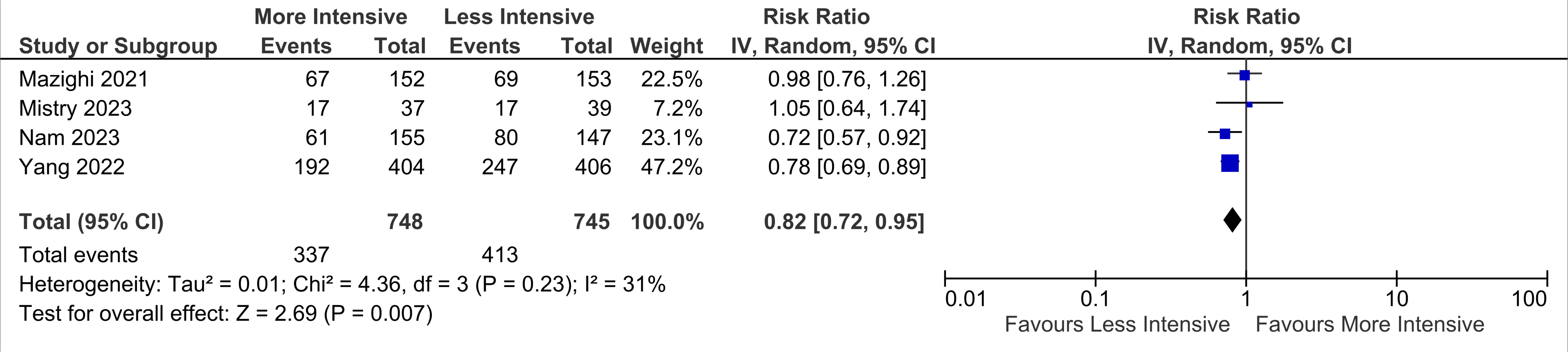
Comparison of participants showing functional independence (modified Rankin scale mRS score=0-2) at 90 days between more intensive blood pressure control and less intensive blood pressure control.

**Figure 3.**
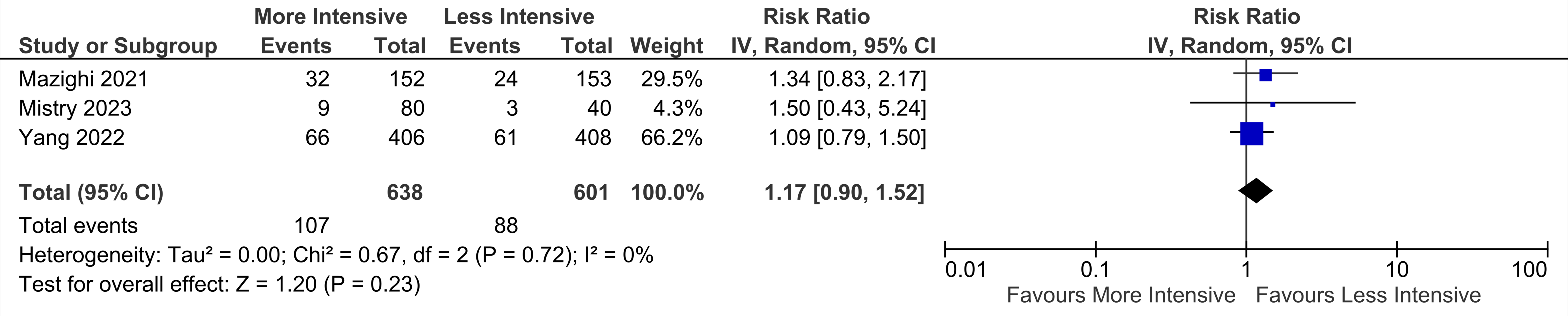
Comparison of 90-day mortality between more intensive blood pressure control and less intensive blood pressure control.

**Figure 4.**
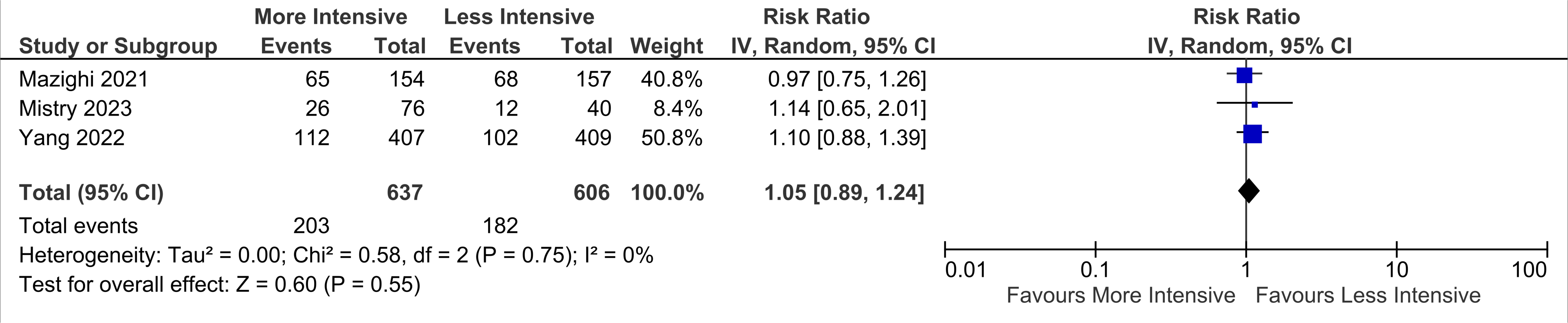
Comparison of incidence of intracerebral hemorrhage (ICH) between more intensive blood pressure control and less intensive blood pressure control.

**Figure 5.**
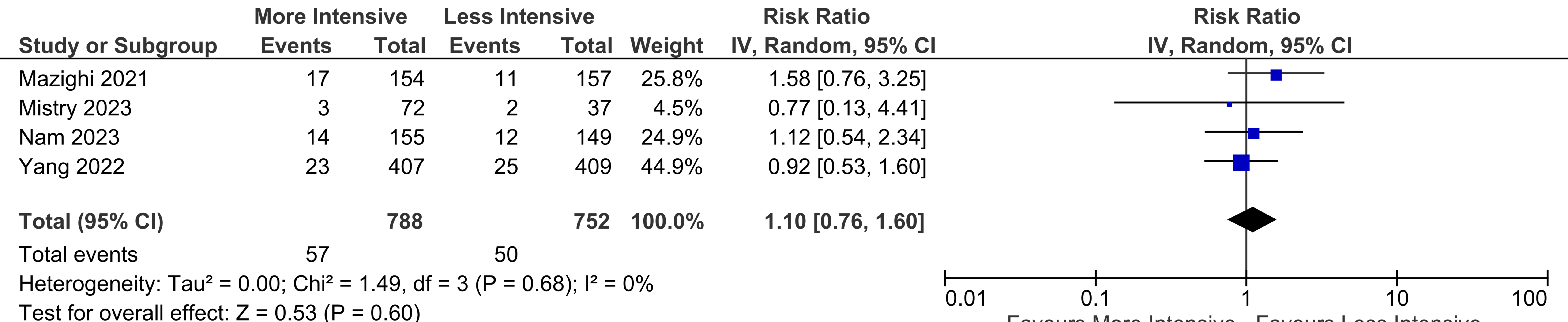
Comparison of incidence of symptomatic intracerebral hemorrhage (sICH) between more intensive blood pressure control and less intensive blood pressure control.

After the study selection, two reviewers extracted the subsequent data into a structured Excel spreadsheet using a pre-piloted form. The extracted information included study characteristics, participant details, intervention specifics, and outcome measures. A third reviewer resolved any discrepancy.

#### Risk of bias assessment

The risk of bias in the included studies was assessed using the revised Cochrane “Risk of bias” tool for randomized trials (RoB 2.0). Two reviewers independently evaluated five specific bias domains, resolving disagreements through discussion or involving a third reviewer if necessary. They graded each included study as low, high, or some concerns regarding bias.

#### Data Synthesis

We reported dichotomous outcomes as relative risk (RR) and 95% confidence intervals. We used the DerSimonian and Laird random-effects model in our meta-analyses. We calculated the Chi^2^ test and *I^2^* statistic to detect and quantify heterogeneity. We interpreted *I^2^* values according to the Cochrane Handbook for Systematic Reviews of Interventions, section 10.10. Regarding interpreting *I^2^* values, 0-40% might not be important, 30-60% may represent moderate heterogeneity, 50-90% may signify substantial heterogeneity, and 75-100% accounts for considerable heterogeneity. P < 0.10 was considered statistically significant for the Chi2 test. All statistical analyses were performed using Review Manager (RevMan, Version 5.4; The Cochrane Collection, Copenhagen, Denmark). The study characteristics and findings of the included studies were presented as tables.

## Results

### Study selection and characteristics of included studies

After applying the eligibility criteria, four RCTs comprising 1560 patients ^17,18,22,23^ were included in our meta-analysis. The age range of participants in all four studies was 68-82 years. Blood pressure targets varied slightly among different studies and are presented in Table 1. The study characteristics of each study are displayed in Table 1.

**Table 1.** Characteristics of included studies.

| Study ID | Yang 2022 | Mazighi 2021 | Nam 2023 | Mistry 2023 |
| --- | --- | --- | --- | --- |
| Country | China | France | South Korea | US |
| Mean Age (years) | 68 ± 4.9 vs 67 ± 4.9 | 76.25 ± 5.5 vs 74 ± 5.8 | 73.2 ± 12.1 vs 72.9 ± 10.8 | 74.9 ± 5.2 vs 69.7 ± 4.0 vs 67.6 ± 5.3 |
| Male n (%) | 249 (61%) vs 257 (63%) | 81/158 (51%) vs 72/160 (45%) | 92(59.4%) vs 88(59.9%) | 12(30%) vs 19(47.5%) vs 20(50%) |
| Sample Size | 407 + 409= 816 | 158 + 160 =318 | 155 + 151=306 | 40 + 40+ 40 (120) |
| Blood Pressure Targets | SBP: 120 vs140–180 | 100-129 vs 130-185 | SBP:<140 vs 140-180 | <140 vs <160 vs <=180 |
| Past History of Hypertension n (%) | 267 (66%) vs 261 (63.8%) | 72/160 (45%) vs 113/160 (71%) | 121(78.1%) vs 110(74.8%) | 32(80%) vs 28(70%) vs 32(80%) |
| Past History of Diabetes n (%) | 81 (19.9%) vs 82 (20%) | 34/155 (22%) vs 33/159 (21%) | 65(41.9%) vs | 12(30%) vs 15(37.5%) vs |
|  |  |  | 62(42.2%) | 13(32.5%) |
| **Past History of Hyperlipidemia n (%)** | 14 (3.4%) vs 13 (3.2%) | 59/153 (39%) vs 55/158 (35%) | 61(39.4%) vs 54(36.7%) | 33(82.5%) vs 28(70%) vs 34(85%) |
| **Baseline antiplatelet use n (%)** | 34 (8%) vs 39 (10%) | 44/156 (28%) vs 37/160 (23%) | NA | 14(35%) vs 13(32.5%) vs 19(47.5%) |
| **Baseline anticoagulant use n (%)** | 20 (5%) vs 20 (5%) | 36/156 (23%) vs 34/160 (21%) | NA | 10(25%) vs 3(7.5%) vs 9(22.5%) |
| **Mean NIHSS score** | 15 vs 15 | 18 (12–20) vs 17 (13–20) | NIHSS score:<br>0-5:14 vs18<br>6-15: 83 vs 89<br>>=16: 58 vs 50 |  |
| **mTICI score 2b n (%)** | 37 (9%) vs 43 (11%) | 70/158 (44%) vs 76/160 (48%) | NA | 16 (40%) vs 17 (42.5%) vs 17 (42.5%) |
| **mTICI score 2c n (%)** | 28 (7%) vs 26 (6%) | NA | NA | 6 (15%) vs 5 (12.5%) vs 7 |
|  |  |  |  | (17.5%) |
| **mTICI score 3 n (%)** | 342 (84%) vs<br>340 (83%) | 88/158 (56%) vs<br>84/160 (52%) | NA | 18 (45%) vs 18<br>(45%) vs 16 (40%) |
| **Occlusion site**<br>**Isolated MCA** | NA | 117/158 (74%) vs<br>119/158 (75%) | NA | M1= first segment<br>of MCA; M2=<br>second segment of<br>MCA<br>M1 31 (77.5%) vs<br>25 (62.5%) vs 23<br>(57.5%)<br>M2 6 (15%) vs 8<br>(20%) vs 15<br>(37.5%) |
| **Occlusion site**<br>**Tandem MCA or**<br>**ICA** | M1 segment of<br>the middle<br>cerebral artery<br>(310 [48%] of<br>643 patients | 41/158 (26%) vs<br>39/158 (25%) | NA | ICA:- 7 (17.5%) vs<br>9 (22.5%) vs 5<br>(12.5%) |

The risk of bias was found to be low in all RCTs except two trials. The study by Mazighi et al. ^23^ had a high risk of bias due to concerns regarding the randomization process and deviations from intended interventions. The study by Mistry et al. also demonstrated a high risk of bias due to concerns regarding the randomization process. The risk of bias of individual trials is presented in Supplementary Figure 1.

### Outcomes

#### Incidence of 90-Day mRS= 0–2 score

In our analysis, we found that more intensive blood pressure management was associated with a statistically significant decrease in patients showing functional independence (modified Rankin scale mRS score=0-2) at 90 days (RR 0.81; CI = 0.72-0.91). There was minimal statistical heterogeneity (*I^2^* = 12%) among different studies.

#### Incidence of 90-day mortality

Our pooled results showed no statistically significant difference between the more intensive blood pressure management group and the less intensive blood pressure management group (RR 1.17; CI = 0.90-1.52). The heterogeneity was minimal among different studies (*I^2^*= 0%).

#### Incidence of any ICH

There was no statistically significant difference between the two groups for the incidence of intracerebral hemorrhage (ICH) (RR 1.05; CI = 0.90-1.23). The heterogeneity was calculated to be 0%.

#### Incidence of Symptomatic ICH

There was no statistically significant difference between the two groups for the incidence of symptomatic intracerebral hemorrhage (sICH) (RR 1.10; CI = 0.76-1.60; *I^2^* = 0%).

## Discussion

In our metanalysis of four RCTs with 1560 patients, we assessed the safety and efficacy of intensive blood pressure management compared to conservative blood pressure management in patients who underwent endovascular thrombectomy for acute ischemic stroke. Our analysis showed that intensive blood pressure lowering is comparable to conservative management.Patients showed a significant decline in functional independence (mRS score 0-2) after intensive blood pressure lowering. There was no statistically significant difference between the intensive blood pressure lowering treatment group and the conventional blood pressure management group regarding 90-day mortality^17,18,22,23^. Furthermore, lowering intensive blood pressure had no significant effect on the incidence of intracerebral hemorrhage and was equivalent to conventional management^17,18,22,23^.

Our comparison parallels the results of prior meta-analysis ^16^, which reports that intensive blood pressure lowering is equivalent to, or less favorable than, conventional BP lowering after successful endovascular thrombectomy for acute ischemic strokes. Previously, the functional independence at 90 days was significantly better in the conventional BP lowering group (SBP<140mmHg) as compared to the intensive group (SBP<130mmHg) ^16^. Zhou et al. ^16^ reported no statistically significant difference in 90-day mortality between the two BP-lowering groups. These results are in agreement with our pooled analysis. Regarding the incidence of ICH, they reported that intensive BP lowering (SBP<140mmHg) was associated with better outcomes, i.e., there was a statistically significant decrease in the incidence of symptomatic ICH as compared to conventional management (SBP <120 mm Hg) ^16^. However, our analysis showed no statistically significant difference in the incidence of sICH between intensive BP lowering and worse functional outcomes. These dissimilar findings could be explained by the overall lower risk of bias in our study, as we included only RCTs in our analysis. It could also be explained by the inclusion of the two large-scale RCTs in our study.

The review encompasses research completed across diverse resource settings and ethnic communities, augmenting our findings’ generalizability. Our study reduced bias by including only RCTs and increased overall power by combining the results of four RCTs, which involved 1560 patients.

It is imperative to take into account the limitations of our analysis. Despite pooling a large cumulative sample size, our meta-analysis was still underpowered for most clinical outcomes assessed, as indicated in the results. Some RCTs had different cutoffs for intensive blood pressure than other studies. Another limitation of the study was that we evaluated study-level data instead of patient-level data, which is a better data source. Some of the included RCTs had flaws with randomization, blinding, and the balance of prognostic factors. Furthermore, one of the RCTs included in the analysis was an open-labeled study^22^, thus imparting some bias in our results.

Our findings suggest intensive blood pressure control following a successful EVT in an acute ischemic stroke should be avoided. Despite the similarity of the results between the RCTs, the conclusion cannot be generalized as only a few studies have been conducted on this topic, and the effect of lowering blood pressure needs to be explored further.

Further studies in the future should be aimed at identifying individual blood pressure-lowering drugs and their effects on mortality. Future RCTs should also be conducted to determine the impact of other interventions, such as early initiation of dual antiplatelet and lipid-lowering agents, in lowering post-stroke mortality following thrombectomy.

## Conclusion

According to our meta-analysis, no benefit of intensive lowering of blood pressure was observed in terms of functional independence at 90 days, mortality rates, and incidence of intracerebral hemorrhage. Future large-scale trials should focus on other interventions to improve prognosis in these patients.

## Statements and Declarations

### Declarations of interest

The authors declare that they have no conflicts of interest and no financial interests related to the material of this manuscript.

### Funding

No financial support was received for this study.

### Ethics approval

No ethical approval was required for this study.

### Consent

No consent was required for this study.

## Data Availability

All data produced in the present work are contained in the manuscript.

## Acknowledgments

N/A.

## Notes

### Competing Interest Statement

The authors have declared no competing interest.

### Funding Statement

This study did not receive any funding.

### Author Declarations

The study used only openly available human data and all the data has been published in different journals and referenced in the manuscript.

