## Supplementary Figure 1 for "Intensive Versus Conservative Blood Pressure Lowering after Endovascular Therapy in Stroke: a meta-analysis of randomized controlled trials"

**Authors’ affiliations:**

**^1.^** Department of Medicine, Cardiff University School of Medicine, Cardiff, United Kingdom

**^2^**^.^ Department of Medicine, Kasturba Medical College, Manipal, India

**^3.^** Royal Lancaster Infirmary, United Kingdom

**^4.^** Department of Medicine, Avalon University School of Medicine, Curaçao

**^5.^** Universidad CES, Medellin, Colombia

**^6^**^.^ Department of Medicine, Sri Ramachandra University, India

**^7.^** Department of Medicine, King Edward Medical College, Pakistan

**^8.^** Department of Medicine, B.J. Medical College, Pune, India

**^9.^** Department of Neurology, University of Alabama

**^10.^** Department of Cardiology, Lahey Hospital and Medical Center, Burlington, MA

**Keywords:** meta-analysis; stroke; endovascular therapy

***Corresponding Author:**

Muhammad Ayyan

**Address:** Department of Medicine, King Edward Medical University, Nila Gumbad Chowk, Neela Gumbad Lahore, Punjab, Pakistan 54000

**ORCID:** 0000-0002-4023-7956 **Twitter handle:** @ayyan_77

**Supplementary Figure 1. Risk of bias of individual studies using the revised Cochrane "Risk of bias" tool for randomized trials (RoB 2.0)**

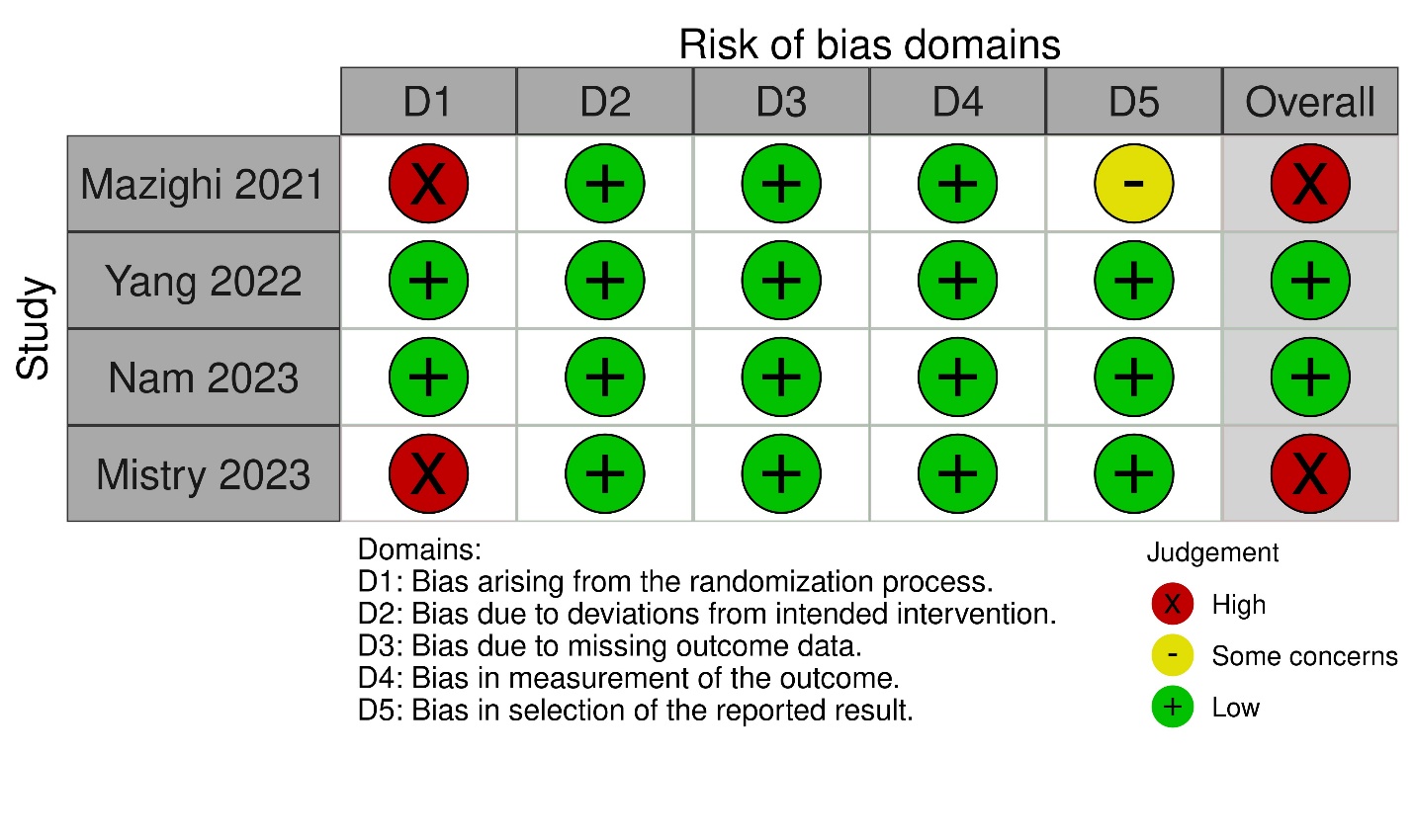
